# Tuberculosis Following In Vitro Fertilization: A Systematic Review of Case Reports, Case Series, and Cohort Studies

**DOI:** 10.1101/2024.10.20.24315846

**Authors:** Ravindra Kumar Garg, Hardeep Singh Malhotra, Vinay Suresh, Rajiv Garg, Amita Jain, Sarika Gupta, Smriti Agrawal

## Abstract

**Introduction:** Tuberculosis (TB) remains a leading global health threat, with increasing concerns regarding post-in vitro fertilization (IVF) TB, especially in high-burden areas. Post-IVF maternal TB poses serious risks to both pregnant women and fetuses, often leading to miscarriage, fetal malformation, or mortality. Congenital TB, though rare, can result from maternal infection, with potentially devastating outcomes.

**Methods:** A systematic review of case reports, case series, and cohort studies was conducted following PRISMA guidelines. Databases including PubMed, Scopus, Embase, and Google Scholar were searched until October 7, 2024, for studies related to TB following in vitro fertilization. Cases were evaluated for clinical presentations, diagnostics, treatment, and outcomes. We separately reviewed 11 cohort studies due to the lack of available individual patient data and the considerable variability in their methodologies.

**Results:** Thirty-seven reports comprising 48 cases were analyzed, with the majority from Asia. Post-IVF TB is commonly presented as miliary TB in mothers, with systemic involvement including pulmonary, central nervous system (CNS), and genitourinary manifestations. Newborns frequently develop congenital TB, presenting with respiratory distress and neurological issues. The diagnosis was confirmed through techniques such as sputum polymerase chain reaction (PCR), endometrial biopsies, and imaging. Maternal outcomes improved with anti-TB therapy, while neonatal outcomes varied, with high mortality rates.

**Conclusion:** Post-IVF TB manifests in mothers as miliary TB with systemic involvement; congenital TB in newborns often leads to respiratory distress, neurological complications, and high mortality. TB screening before IVF is critical in high-risk regions to prevent severe maternal and neonatal complications.

## Introduction

In 2022, tuberculosis (TB) was responsible for 1.3 million deaths, including 167,000 among individuals living with HIV, making it the second deadliest infectious disease after COVID-19. Approximately 10.6 million people developed TB, with men comprising 5.8 million cases, women 3.5 million, and children 1.3 million. Eight countries, including India, Indonesia, and China, represented almost two-thirds of the global TB burden. The South-East Asia region had the highest number of cases (46%), followed by Africa (23%) and the Western Pacific (18%). Multidrug-resistant TB remains a significant challenge, with only 40 percent of the 410,000 drug-resistant cases receiving treatment.^**1**^

Post-in vitro fertilization (IVF) maternal TB is a growing concern, particularly as in vitro fertilization becomes more widely used for infertility treatment. TB, especially in its military form, poses serious risks to both pregnant women and fetuses, often leading to miscarriage, fetal malformation, or even maternal mortality. During pregnancy, hormonal changes, such as increased estrogen levels, can weaken the immune system, increasing the risk of TB reactivation. The incidence of TB during pregnancy has risen with the growing application of in vitro fertilization, especially in regions with high TB prevalence.^**2**,**3**^

Congenital TB is rare, but newborns are at risk for rapid progression if infected. Congenital TB is a rare condition, it occurs when the infection is transmitted from the mother to the fetus, typically through the placenta. With the increasing use of in vitro fertilization, particularly in women with latent or undiagnosed genital TB, congenital TB has become more prevalent in developing countries. Despite the rarity of congenital TB, the lack of routine TB screening before in vitro fertilization in high-burden regions poses significant risks to newborns, especially in cases of untreated or reactivated maternal TB.^**4**,**5**^

The objective of this systematic review is to assess the spectrum of TB in women and newborns following in vitro fertilization, focusing on the impact of maternal TB on pregnancy and newborn outcomes, particularly congenital TB, through analysis of case reports, case series, and cohort studies.

## Methods

We conducted a systematic review of case reports, case series, and cohort studies on TB following IVF, following the PRISMA guidelines for systematic reviews. The protocol for this review is registered with PROSPERO under the registration number CRD42024597967.^**6**^

### Search strategy

Our systematic review included thorough searches across several databases such as PubMed, Scopus, Embase, and Google Scholar. The search in Google Scholar was limited to the first 50 pages of results. Non-English articles were translated using Google Translate. A specific search string was utilized to retrieve relevant studies.

We used the search string “((“In Vitro Fertilization” OR IVF OR “Assisted Reproductive Technology” OR ART OR “Fertility Treatment” OR “Reproductive Techniques” OR “Embryo Transfer” OR “Ovulation Induction” OR “Egg Retrieval”) AND (Tuberculosis OR TB OR “*Mycobacterium tuberculosis*” OR “Pulmonary Tuberculosis” OR “Extrapulmonary Tuberculosis” OR “Genital Tuberculosis” OR “Urogenital Tuberculosis” OR “Tuberculous Infection”)”. The last search was undertaken on October 7, 2024.

### Definitions

#### Miliary TB

Miliary TB is characterized on chest radiographs by tiny, uniformly sized pulmonary opacities, each measuring 2 mm or less in diameter, widely distributed throughout the lungs, resembling millet seeds.

#### Disseminated TB

Disseminated TB is defined as the involvement of two or more non-adjacent body sites caused by the spread of *Mycobacterium tuberculosis*.

#### Congenital TB

Cantwell’s criteria for diagnosing congenital tuberculosis require confirmation of tuberculous lesions in the newborn. Additional criteria include the appearance of lesions within the first week of life, the presence of a primary hepatic complex or caseating granulomas in the liver, and evidence of TB in the placenta or maternal genital tract. To exclude postnatal transmission, the infant must be separated from potential sources of infection, and all contacts should be thoroughly examined.^**7**^

### Study Selection And Data Extraction

The evaluation process occurred in two distinct phases. First, two reviewers (RKG and VS) independently reviewed the titles and abstracts, followed by a comprehensive examination of the full-text articles that met the inclusion criteria. In cases of disagreement, a third reviewer (HSM) intervened to resolve them. A different pair of reviewers (SA and SG) then assessed the quality of the included studies, and any disputes were addressed through discussion, with a third opinion sought if necessary. Records that lacked adequate information were excluded from the final analysis.

EndNote 21 software (Clarivate Analytics, Philadelphia, PA, USA) was utilized to manage and detect duplicate records, with oversight from two assessors (RKG and HSM). A third assessor (SG) was available to resolve any discrepancies. The selection process at each stage was illustrated in a PRISMA flowchart, generated using EndNote 21.

Data were gathered on patient demographics, clinical presentations, duration of illness, maternal and neonatal TB following IVF manifestations, diagnostic methods, treatment protocols, and outcomes. Three reviewers (RG, VS, SG) conducted the data extraction, with a fourth (RKG) mediating any disputes. The systematic review synthesized the information into comprehensive tables, supported by relevant figures and statistical analyses, using Microsoft Excel for data analysis. The final report provided a detailed summary of TB cases following IVF, drawing from case reports, case series, and cohort studies.

### Quality assessment

Each case was carefully assessed based on four essential parameters: selection, validation, causality, and reporting, following the framework provided by Murad MH et al.^**8**^ Cases were classified according to quality, with “good quality” assigned to those meeting all criteria as per Della Gatta et al.’s guidelines. Cases that fulfilled three criteria were considered “fair quality,” while those meeting only one or two criteria were deemed “poor quality.”^**9**^ we did not perform the quality assessment for cohort studies.

### Data Synthesis

The data were meticulously gathered and compiled into a Word document, emphasizing key details. Both narrative and qualitative approaches were employed for data synthesis. Categorical data were expressed as frequencies and percentages. This information was methodically structured into tables within the final report for the systematic review of TB following IVF, encompassing case reports, case series, and cohort studies.

## Results

A total of 37 reports comprising 48 cases of TB following IVF were analyzed. Among these, 41 cases involved pregnant women, and 9 involved newborns. All the cases were classified as having good or fair quality (Supplementary item-1). The PRISMA checklist has been provided as supplementary item 2. Patient-related data extracted from 37 reports have been compiled in supplementary item 3.

The majority of cases were reported from China (n=13), followed by the United States (n=5) and Turkey (n=3). These reports were distributed across four continents, with Asia reporting the highest number (n=20), followed by Europe (n=9), North America (n=7) and 1 from Australia. We separately reviewed 11 cohort studies due to the lack of available individual patient data and the considerable variability in their methodologies.

The duration of illness in patients varied widely, from immediate onset at birth or post-delivery to up to 60 days postpartum. Symptoms appeared in some cases within 3 to 5 days, while others took longer, such as 28 or 56 days. Gestational factors influenced symptom onset, with some cases emerging as early as the 9th week or after premature delivery. In several instances, illness duration was not reported, while specific timeframes like 14, 30, or 49 days were observed in others.

Post-IVF TB in mothers and newborns presents a wide range of symptoms. In mothers, common signs include fever, cough, headache, and respiratory insufficiency, often progressing to more severe issues like seizures, coma, vomiting, acute respiratory distress syndrome (ARDS), vision problems, and vaginal bleeding. Severe systemic involvement may lead to respiratory failure, septic shock, and hepatosplenomegaly. These symptoms often overlap with typical systemic and respiratory TB, intensified by pregnancy and IVF-related factors.

For newborns, respiratory distress is a primary concern, often occurring soon after birth. Other critical symptoms include late-onset sepsis, fever, pulmonary infiltrates, hepatosplenomegaly, and neurological issues such as seizures. Respiratory failure is also common, reflecting the severity of the infection. Both mothers and newborns frequently show overlapping systemic and respiratory signs.

Post-IVF TB in pregnant mothers primarily presented as miliary TB, often involving pulmonary lesions with chest computed tomography (CT) or X-ray showing miliary nodules or ground-glass opacities. Disseminated TB affecting organs such as the liver, placenta, eyes, and CNS was also common, with some mothers presenting with central nervous system (CNS) TB, evident by tuberculomas or meningoencephalitis on brain magnetic resonance imaging (MRI). Genitourinary TB, endometrial TB, and complications like cold abscesses or chronic ascites were noted in several cases. Pre-existing TB, including calcified nodules and fibrotic lung changes, were also detected in some mothers through X-rays or other diagnostic methods.

In newborns, congenital TB was frequently observed, often with disseminated disease affecting multiple organs. Chest X-rays and CT scans revealed bilateral pneumonia and miliary TB. Some newborns had multi-drug resistant TB, resistant to key drugs like isoniazid and rifampicin, with involvement of organs such as the liver, spleen, and heart. CNS TB was also detected in several newborns, along with mediastinal lymphadenopathy causing tracheal and bronchial compression. Common findings included pneumonia, consolidation, cavitation, and calcifications in the lungs.

Microbiological confirmation of TB in both mothers and newborns was achieved using several diagnostic techniques. *Mycobacterium tuberculosis* was identified through sputum polymerase chain reaction (PCR), GeneXpert, gastric aspirates, and blood mNGS. In mothers, endometrial and liver biopsies showed necrotizing granulomas, while CSF analysis was crucial for diagnosing CNS TB, with positive PCR results and typical TB meningitis markers. In newborns, *Mycobacterium tuberculosis* was confirmed through endotracheal aspirates, gastric aspirates, and CSF samples.

Maternal outcomes were generally positive, with most mothers improving significantly after treatment with anti-TB therapy, and some fully recovering within months of follow-up. For example, in China, mothers showed marked improvement within 3 to 6 months of treatment. Newborn outcomes, however, were more variable.

Premature deliveries were common, and the mortality rate was high, with nearly 30% of cases resulting in neonatal death or spontaneous abortion. For instance, a newborn in China born at 27 weeks did not survive despite intensive care. On the other hand, some newborns made full recoveries with intensive treatment, such as a surviving twin in Italy who improved after 15 months of treatment.

### Analysis of cohort studies

We also analyzed 11 cohort studies, where individual patient data were not available

### Post-IVF TB in Pregnant Women

Several studies analyzed cases of TB in pregnant women who conceived through IVF-ET. These women typically experienced more severe TB complications compared to those with natural pregnancies. Common symptoms included early-onset fever, dyspnea, and cough. Miliary TB was frequently observed, with many cases also involving extrapulmonary forms such as tuberculous meningitis, peritonitis, and pleurisy. Imaging often revealed diffuse miliary nodules, ground-glass opacity, and pleural effusion. Some patients developed respiratory failure, with a small number requiring mechanical ventilation or extracorporeal membrane oxygenation (ECMO). IVF pregnancies were associated with high rates of adverse pregnancy outcomes, including spontaneous and inevitable abortions, preterm labor, and neonatal death. One study reported that 60% of IVF pregnancies resulted in miscarriage or preterm labor, and only one baby survived. Complications like vaginal bleeding and maternal criticality were more frequent in IVF patients, indicating a more severe disease course. Despite anti-TB treatment, these women often experienced poor pregnancy outcomes, emphasizing the need for TB screening and preventive treatment before undergoing IVF.

### Congenital TB

Congenital TB was analyzed in children born to mothers with TB, particularly those conceived through IVF. In most cases, symptom onset in infants occurs within the first four weeks of life. These children, especially those born after IVF, had more severe TB forms, including tuberculous meningitis and liver TB. Common clinical presentations included fever, respiratory distress, and seizures. Imaging studies frequently showed pulmonary nodules and evidence of disseminated TB. Diagnostic tools such as T-SPOT.TB was helpful, with high positivity rates, although cerebrospinal fluid cultures had lower positivity rates. Drug-resistant TB was rare, and anti-TB treatment often included isoniazid and rifampicin. However, children born to IVF mothers had poorer outcomes, including higher rates of mortality, liver injury, and developmental delays. Early diagnosis, maternal screening, and proper management of latent or active TB in mothers are essential to improve outcomes for both mothers and infants in these high-risk pregnancies.

## Discussion

This systematic review analyzed 37 reports comprising 48 cases of TB following in vitro fertilization. TB cases were predominantly observed in Asia, with a significant portion affecting both mothers and newborns. Post-IVF TB is commonly presented as miliary TB with systemic involvement, including pulmonary, central nervous system, and genitourinary manifestations. Newborns often develop congenital TB with respiratory distress, pneumonia, and neurological issues. Diagnostic confirmation was achieved using techniques like sputum PCR and endometrial biopsies. Maternal outcomes improved with anti-TB therapy, but newborn outcomes varied, with a high mortality rate and frequent premature deliveries. These findings emphasize the importance of TB screening before IVF.

TB negatively affects pregnancy outcomes, increasing risks of maternal morbidity, preterm delivery, low birth weight, and neonatal complications. The presence of HIV further raises the risk of active TB, especially in the postpartum period. Extrapulmonary TB, particularly in the central nervous system, presents severe risks with delayed diagnosis often leading to poor pregnancy and fetal outcomes. Miliary TB has emerged in women undergoing in vitro fertilization, sometimes causing respiratory failure or acute respiratory distress syndrome. Given these risks, screening for TB before fertility treatments is crucial, especially in regions with high TB incidence. Initiating anti-TB therapy before IVF can help improve pregnancy outcomes.^**10**^

Post-IVF maternal TB likely results from a combination of immune suppression, hormonal changes, and reactivation of latent TB. Pregnancy naturally suppresses cell-mediated immunity, crucial for controlling *Mycobacterium tuberculosis*. Elevated estrogen and progesterone levels during pregnancy and controlled ovarian hyperstimulation for IVF further impair T-cell function, increasing vulnerability to reactivation. Studies suggest latent TB in the reproductive organs can be reactivated due to these hormonal and immune changes. Additionally, invasive procedures like egg retrieval may trigger local inflammation, facilitating TB reactivation. The rapid hormonal shifts during IVF may exacerbate this risk, especially in areas with a high latent TB burden.^**11**^ In women with untreated genital TB, IVF poses significant risks, including reactivation of TB, embryo implantation failure, and increased miscarriage rates. Administering antiTB treatment before starting IVF can improve pregnancy outcomes by enhancing ovarian reserve and endometrial receptivity.^**12**^ Studies reveal latent TB infection prevalence in pregnancy, especially in higher-burden regions like South Africa and India is quite high. Cytokine expression, such as interferon-inducible protein 10, varies with gestational age, potentially serving as biomarkers for TB in pregnancy, particularly in the third trimester.^**13**^ A study followed 101 mother-infant pairs, with 23 infants (23%) diagnosed with TB. A positive maternal sputum culture at delivery significantly increased the risk of TB in infants.

Contributing factors included poor maternal treatment adherence, health system failures, and non-maternal transmission.^**14**^ During pregnancy, hormonal and immune system changes reduce T lymphocyte activity, making individuals more vulnerable to TB infection and reactivation. IVF pregnancies, which are associated with increased levels of progesterone and corticosteroids, further elevate this risk. Latent TB and genital TB, which are frequent causes of infertility in women, often go undiagnosed, increasing the likelihood of reactivation during pregnancy.^**15**^

Post-IVF congenital TB occurs when *Mycobacterium tuberculosis* is transmitted from the mother to the fetus, typically due to the reactivation of latent TB during pregnancy. Hormonal changes associated with IVF, particularly increased estrogen and progesterone, suppress maternal immunity, allowing latent TB to reactivate. The bacteria can then spread hematogenously to the placenta and cross into the fetal circulation, leading to transplacental transmission. The fetus may also be exposed through infected amniotic fluid. Given the fetus’s immature immune system, *Mycobacterium tuberculosis* can disseminate to vital organs, causing severe conditions like miliary TB or tuberculous meningitis, often with devastating outcomes post-delivery. A review analyzed 22 cases of congenital TB in infants conceived through in vitro fertilization, highlighting that most mothers were from countries with high TB incidence. Nearly all infants were born prematurely, with TB symptoms typically appearing around 28 days of life. Pulmonary TB was common, while some cases involved disseminated disease. Many mothers had undiagnosed or untreated genital TB, a known cause of infertility, particularly in high TB-burden regions. The findings emphasize the importance of TB screening before fertility treatments, especially in women from endemic areas, to reduce the risk of congenital TB in infants.^**16**^

The study’s limitations include a small sample size, retrospective design, data heterogeneity, and limited long-term follow-up. Moreover, diagnostic variability, selection bias, and a geographic focus on high TB prevalence areas affect its generalizability. The extensive data heterogeneity made it challenging to count frequencies and calculate percentages.

In conclusion, post-IVF TB presents significant risks for both mothers and newborns. Early screening, diagnosis, and treatment are crucial to improving maternal and neonatal outcomes, especially in TB-endemic regions.

**Figure 1.**
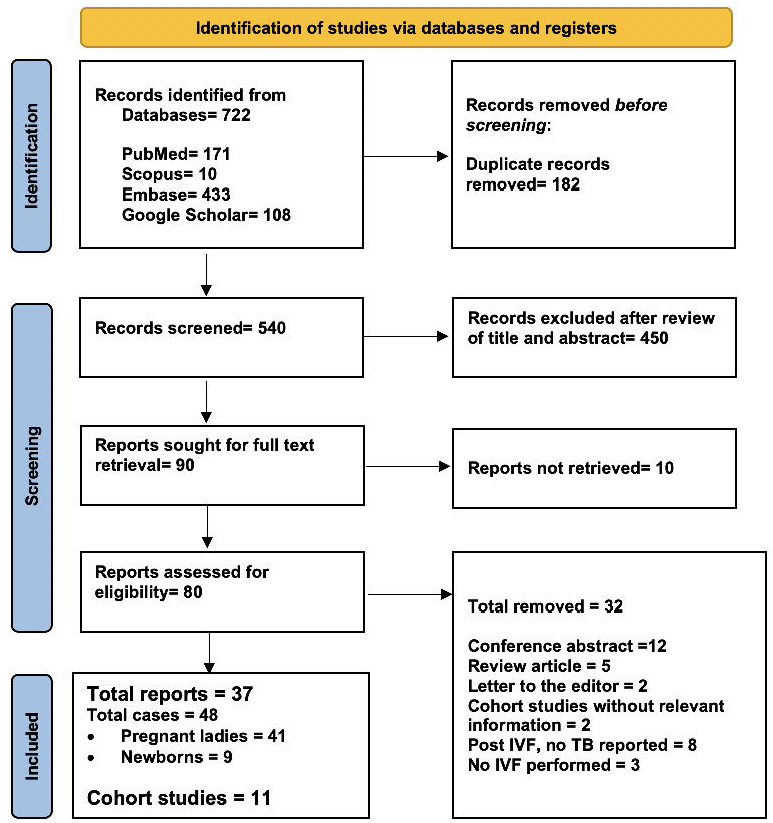
PRISMA flow diagram depicts the case selection process. (TB= Tuberculosis; IVF= In vitro fertilization)

## Supporting information

Supplementary item-1

Supplementary item-2

Supplementary item-3

## Declarations

### Ethical Statement

As this study did not involve human or animal subjects, ethical approval was not necessary.

### Data Availability Declaration

All data relevant to this study is included within the manuscript and its supplementary materials.

