## Supplementary item-2 for "Tuberculosis Following In Vitro Fertilization: A Systematic Review of Case Reports, Case Series, and Cohort Studies"

**Supplementary table-2: Evaluation of the methodological quality of case reports and case series
Tuberculosis Following In Vitro Fertilization: A Systematic Review of Case Reports, Case Series, and Cohort Studies**

| **Reference** | **Does the patient represent the whole experience of the investigator** | **Was the exposure adequately ascertained?** | **Was the outcome adequately ascertained?** | **Were other alternative causes that may explain the observation ruled out?** | **Was there a challenge and/or re-challenge phenomenon?** | **Was there a dose-response effect?** | **Was follow-up long enough for outcomes to occur?** | **Is the case(s) described with sufficient details to allow practitioners make inferences related to their own practice?** | **Score** |
| --- | --- | --- | --- | --- | --- | --- | --- | --- | --- |
| Zhang et al 2024 | Yes | Yes | Yes | Yes | NA | Yes | Yes | Yes | 7 |
| Singh et al 2024 | Yes | Yes | Yes | Yes | NA | Yes | Yes | Yes | 7 |
|  | Yes | Yes | Yes | Yes | NA | Yes | NA | NA | 5 |
|  | Yes | Yes | Yes | Yes | NA | Yes | NA | NA | 5 |
| Jiménez-Fuentes et al 2024 | Yes | Yes | Yes | Yes | NA | Yes | Yes | Yes | 7 |
| Campos-Herrero 2024 | Yes | Yes | Yes | Yes | NA | Yes | Yes | Yes | 7 |
|  | Yes | Yes | Yes | Yes | NA | Yes | Yes | Yes | 7 |
|  | Yes | Yes | Yes | Yes | NA | Yes | Yes | Yes | 7 |
| Sands et al 2023 | Yes | Yes | Yes | Yes | NA | Yes | Yes | Yes | 7 |
| Li et al 2023 | Yes | Yes | Yes | Yes | NA | Yes | NA | Yes | 6 |
| Beshar et al 2023 | Yes | Yes | Yes | Yes | NA | Yes | Yes | Yes | 7 |
| Zhuang et al 2022 | Yes | Yes | Yes | Yes | NA | Yes | Yes | Yes | 7 |
| Yue et al 2022 | Yes | Yes | Yes | Yes | NA | Yes | Yes | Yes | 7 |
| Vempati et al 2022 | Yes | Yes | Yes | Yes | NA | Yes | Yes | Yes | 7 |
| Shi and Sun 2022 | Yes | Yes | Yes | Yes | NA | Yes | Yes | Yes | 7 |
|  | Yes | Yes | Yes | Yes | NA | Yes | Yes | Yes | 7 |
| Liu et al 2022 | Yes | Yes | Yes | Yes | NA | Yes | Yes | Yes | 7 |
| Kestens et al 2022 | Yes | Yes | Yes | Yes | NA | Yes | Yes | Yes | 7 |
| Sotskiy et al 2021 | Yes | Yes | Yes | Yes | NA | Yes | Yes | Yes | 7 |
| Ma et al 2021 | Yes | Yes | Yes | Yes | NA | Yes | Yes | Yes | 7 |
| Cheng et a 2021 | Yes | Yes | Yes | Yes | NA | Yes | Yes | Yes | 7 |
| Regalado et al 2020 | Yes | Yes | Yes | Yes | NA | Yes | Yes | Yes | 7 |
| Fan et al 2020 | Yes | Yes | Yes | Yes | NA | Yes | Yes | Yes | 7 |
| Venturini et al 2019 | Yes | Yes | Yes | Yes | NA | Yes | Yes | Yes | 7 |
| Amin et al 2019 | Yes | Yes | Yes | Yes | NA | Yes | Yes | Yes | 7 |
| Zhang et al 2018 | Yes | Yes | Yes | Yes | NA | Yes | Yes | Yes | 7 |
|  | Yes | Yes | Yes | Yes | NA | Yes | Yes | Yes | 7 |
| Samedi et al 2017 | Yes | Yes | Yes | Yes | NA | Yes | Yes | Yes | 7 |
| Namani et al 2017 | Yes | Yes | Yes | Yes | NA | Yes | Yes | Yes | 7 |
| Parmaksiz et al 2016 | Yes | Yes | Yes | Yes | NA | Yes | Yes | Yes | 7 |
| Emiralioglu et al 2016 | Yes | Yes | Yes | Yes | NA | Yes | Yes | Yes | 7 |
| Hongbo and Li 2015 | Yes | Yes | Yes | Yes | NA | Yes | Yes | Yes | 7 |
| Zheng et al 2014 | Yes | Yes | Yes | Yes | NA | Yes | NA | Yes | 6 |
| Ting-ting and Guo-zhong 2014 | Yes | Yes | Yes | Yes | NA | Yes | NA | Yes | 6 |
| Mony et al 2014 | Yes | Yes | Yes | Yes | NA | Yes | Yes | Yes | 7 |
| Flibotte 2013  A report of five patients. | Yes | Yes | Yes | Yes | NA | Yes | Yes | Yes | 7 |
|  | Yes | Yes | Yes | Yes | NA | Yes | Yes | Yes | 7 |
|  | Yes | Yes | Yes | Yes | NA | Yes | Yes | Yes | 7 |
|  | Yes | Yes | Yes | Yes | NA | Yes | Yes | Yes | 7 |
|  | Yes | Yes | Yes | Yes | NA | Yes | Yes | Yes | 7 |
| Jacquemyn et al 2012 | Yes | Yes | Yes | Yes | NA | Yes | Yes | Yes | 7 |
| Stuart et al 2009 | Yes | Yes | Yes | Yes | NA | Yes | Yes | Yes | 7 |
| Altunhan et al 2009  (Twins) | Yes | Yes | Yes | Yes | NA | Yes | Yes | Yes | 7 |
|  | Yes | Yes | Yes | Yes | NA | Yes | Yes | Yes | 7 |
| Chan et al 2003 | Yes | Yes | Yes | Yes | NA | Yes | Yes | Yes | 7 |
| Kissel et al 1999 | Yes | Yes | Yes | Yes | NA | Yes | Yes | Yes | 7 |
| Muller and Seelos 1994 | Yes | Yes | Yes | Yes | NA | Yes | Yes | Yes | 7 |
| Addis et al 1988 | Yes | Yes | Yes | Yes | NA | Yes | Yes | Yes | 7 |

**Domains Leading explanatory questions**

Selection 1. Does the patient(s) represent(s) the whole experience of the investigator or is the selection method unclear to the extent that other patients with similar presentation may not have been reported?

Ascertainment 2. Was the exposure adequately ascertained?

3. Was the outcome adequately ascertained?

Causality

4. Were other alternative causes that may explain the observation ruled out?

5. Was there a challenge/re-challenge phenomenon?

6. Was there a dose–response effect?

7. Was follow-up long enough for outcomes to occur?

Reporting

8. Is the case(s) described with sufficient details to allow other investigators to replicate the research or to allow practitioners make

inferences related to their own practice?
