## Supplementary item-3 for "Tuberculosis Following In Vitro Fertilization: A Systematic Review of Case Reports, Case Series, and Cohort Studies"

**Table-1: Details of clinical characteristics , treatment outcomes in patients with tuberculosis following In Vitro Fertilization (data were derived from isolated cases described in case reports and case series.**

| **Reference** | **Country** | **Maternal age** | **Newborn Age** | **Duration of illness** | **Clinical presentation** | **Type of maternal TB** | **Type of newborn TB** | **Microbiological confirmation** | **Treatment Given**  **Duration of Anti-TB Treatment** | **Maternal Outcome** | **Newborn outcome** |
| --- | --- | --- | --- | --- | --- | --- | --- | --- | --- | --- | --- |
| Zhang et al 2024 | China | NA | 22 days | 8 days | Fever | NA | Congenital disseminated-TB  Resistant to isoniazid, rifampicin, ethambutol, and streptomycin  *Chest X-ray and CT=bilateral*  *pneumonia lesions*  Other lesions in liver, spleen and heart | Sputum PCR= *Mycobacterium tuberculosis DNA*  Gastric juice GeneXpert = positive, rifampicin resistance  Blood mNGS= *Mycobacterium tuberculosis DNA* | Mechanical ventilation  Linezolid containing regimen | Mother is being treated with ATT | Improved  3 months follow up CT= lesions resolved |
| Singh et al 2024 | India | 28  Pregnancy 18 weeks | NA | 10 days | Respiratory insufficiency | Miliary pulmonary TB | NA | NA | MTP  ATT | Discharged after 9 days | Intrauterine fetal death |
|  |  | 31  Pregnancy 22 weeks  Twin pregnancy | NA | 60 days | Cough  Respiratory insufficiency | Miliary pulmonary TB | NA | NA | ATT | Died | NA |
|  |  | 32  Pregnancy 12 weeks | NA | 15 days | Fever  Respiratory insufficiency | Miliary pulmonary TB | NA | NA | ATT | NA | Premature delivery later baby died. |
| Jiménez-Fuentes et al 2024 | Spain | 41 | Pregnancy 23 weeks | NA | Cough | Miliary pulmonary TB | NA | Gastric aspirate GeneXpert = *Mycobacterium tuberculosis DNA* | ATT 6 months | Improved | The newborn, premature and of low weight |
| Campos-Herrero 2024 | Spain | 30 Pregnancy 24 weeks  Twin born out of caesarean section | NA | NA | Fever, cough  Respiratory insufficiency | Endometrial TB | NA | Endometrial biopsy Ziehl-Neelsen staining= Mycobacterium *tuberculosis* | ATT | Mother improved | Not applicable |
|  |  | Twin 1  Immediately after delivery | NA | Immediately after delivery | Respiratory insufficiency | NA | Congenital TB | Lymph node biopsy and  Gastric aspirate NAAT = Mycobacterium *tuberculosis* | ICU care and ATT | NA | Improved |
|  |  | Twin 2 | NA | Immediately after delivery | Respiratory insufficiency | NA | Congenital TB | Liver granuloma and  Gastric aspirate smear = Mycobacterium *tuberculosis* | ICU care and ATT  Corticosteroids | NA | Improved |
| Sands et al 2023 | USA | NA  Mother was Indian immigrant | 29.6 weeks, born vaginally with a birth weight of 1440 | Immediately after delivery | Respiratory insufficiency | Mother had Genito-urinary TB | Multi-drug resistant congenital TB  X-ray = Lobar pneumonia | Lung biopsy and gastric aspirates= = *Mycobacterium tuberculosis MDR* | ICU care and ATT 18 months | Mother took 18 months ATT and she improved | Improved |
| Li et al 2023  (A report of two patients, one did not have IVF) | China | 33  Did not had endometrial TB | *10 weeks of gestation* | 10 days | Cough  Respiratory insufficiency  Fever | Miliary TB  X-ray and CT chest = *ground-glass opacities*  *and diffuse miliary nodules* | NA | *Alveolar*  *lavage fluid= Mycobacterium tuberculosis DNA* | ATT 12 months  ICU care | Mother improved clinically as well on imaging | NA |
| Beshar et al 2023 | USA | 32  Indian immigrant | NA | 49 days | Severe headache | Disseminated TB  MRI brain = multiple miliary lesions  CT thorax= multiple miliary lesions  PET= intense heterogenous hypermetabolic activity in the endometrium,  cervix, and left external iliac nodes and T10 spinal lesion | NA | CSF= Normal  serum QuantiFERON-TB Gold test was positive  Endometrial biopsy culture= positive | ATT 12 months | Mother improved clinically as well on imaging | Fetal demise at 23 weeks of gestation |
| Zhuang et al 2022 | China | 35 | Infant  *born at 27+5 weeks of gestation* | At birth | Respiratory insufficiency  Skin rash | Mother had endometrial TB | Congenital TB  X-ray= lung pneumonia | *Endotracheal aspirate= Mycobacterium tuberculosis* | ICU care and ATT | NA | Died |
| Yue et al 2022  (Article in Chinese) | China | Infant | NA | 30 days | Fever | Mother had endometrial TB | Congenital disseminated TB  X-ray= Miliary pulmonary TB  CNS TB | Gastric aspirates= = *Mycobacterium tuberculosis* | ATT 6 months | NA | improved |
| Vempati et al 2022 | India | 32  27 weeks of gestational  an intrauterine twin pregnancy | NA | 42 days | Seizures later choreo-athetosis | CNS TB  MRI brain= multiple tuberculoma | NA | MRS= a lipid peak | ATT and epileptic drugs Corticosteroids  ICU care  Premature cesarean section | Improved | New born twins were normal |
| Shi and Sun 2022 | China | 27  IVF 7 months ago  Had 10mg Prednisolone per day for 30 days | NA | NA | Fever, headache and vomiting  Seizures and coma | CNS TB  MRI brain= miliary tuberculoma | NA | CSF= Increased cells and protein  Low glucose | ATT and other symptomatic treatment | Improved | Spontaneous abortion after 4 months |
|  |  | 27  IVF 6 months ago  Had 10mg Prednisolone per day for 20 days | NA | NA | Fever, headache and vomiting  Seizures and coma | CNS TB  MRI brain= miliary tuberculoma | NA | CSF= Increased cells and protein  Low glucose | ATT and other symptomatic treatment | Improved | Spontaneous abortion |
| Liu et al 2022 | China | 30  Uterine pregnancy with twins at 8 weeks | NA | 17 days | Fever  Cough  Acute respiratory distress syndrome | Miliary TB, INH resistant  CT chest = miliary lung lesions | NA | Sputum culture = Isoniazid resistant *Mycobacterium tuberculosis* | ATT 10 months and other symptomatic treatment | Improved | Spontaneous abortion |
| Kestens et al 2022 | Belgium | 35  15th week of a twin pregnancy | NA | NA | Fever, cough, dyspnea | Disseminated TB  Liver, Placenta and eys were having tuberculoma  CT chest = miliary lung lesions | NA | Ziehl-Neelsen stain on liver biopsy= *Mycobacterium tuberculosis*  GeneXpert in sputum= positive | ATT 12 months    Drug-induced hepatotoxicity  Alternative ATT regimen | Improved | Spontaneous abortion |
| Sotskiy et al 2021 | Armenia | 30  IVF was done 56 days earlier  Uterine pregnancy with twins at 8 weeks | NA | 17 days | Fever  Cough  Acute respiratory distress syndrome | Miliary tuberculosis  CT chest = miliary lung lesions | NA | Sputum culture = Isoniazid resistant *Mycobacterium tuberculosis* | ATT 12 months and other symptomatic treatment | Improved | Spontaneous abortion |
| Ma et al 2021 | China | 28 | NA | 7 days  after spontaneous abortion | Fever  Cough  Acute respiratory distress syndrome | CT chest = Miliary lung lesions | NA | Alveolar lavage= The mNGS showed three sequences of M. *tuberculosis* complex later culture was positive | Mechanical ventilation and ATT | Died | Spontaneous abortion |
| Cheng et a 2021 | China | 31  2 years secondary infertility | NA | After premature delivery | Persistent fever  Headache and vomiting | CT chest = miliary lung lesions  CNS tuberculosis  Had disseminated TB | The preterm neonate had congenital TB | NA | ATT 2 years | Improved  In next pregnancy, she had normal delivery. | Died |
| Regalado et al 2020 | Mexico | 29  at 17 weeks of twin pregnancy | NA | NA | Fever and thrombocytopenia | CT chest = miliary lung lesions | NA | Sputum Xpert MTB/RIF= *Mycobacterium tuberculosis* | ATT | Improved | inevitable abortion |
| Fan et al 2020 | China | 28  Progesterone injections were given.  Missed abortion at 9 weeks’ gestation | NA | At the 9th week of gestation | Fever | CT chest = miliary lung lesions | NA | Xpert MTB/RIF vaginal secretions = *Mycobacterium tuberculosis*  Uterine curettage = *Mycobacterium tuberculosis* | ATT | Improved | inevitable abortion |
| Venturini et al 2019 | Italy | NA  Female twin born prematurely | at 27 weeks of gestation | 3 days | Respiratory distress after birth  *4 months of age* | NA | Congenital TB  *Multiple enlarged mediastinal and subpleural lymph nodes* | **Mother’s Placenta=** acid-fast bacilli.  Postmortem examination of twin sister= disseminated TB | ATT 15 months  Initially mechanical ventilation | NA | *Twin sister died 60 days after*  *birth*  Other improved |
| Amin et al 2019 | USA | 32  Nigerian woman  Stillborn delivery of twins | NA | NA | Fever and dysnoea | Disseminated TB  Miliary pulmonary TB and CNS TB | NA | **Liver biopsy**= Caseating granulomas  Endometrial biopsy= *Mycobacterium tuberculosis* | ATT | Improved | Stillborn delivery of twins |
| Zhang et al 2018 | China | NA | 30 weeks' gestation, born via cesarean delivery | 56 days | Progressive respiratory failure and septic shock | Pulmonary TB and TB meningitis diagnosed 5 days before delivery | Congenital TB | Tracheal aspirates= *Mycobacterium tuberculosis* | ATT  Initially mechanical ventilation | NA | Died |
|  |  | NA | 31 weeks' gestation, born via primary cesarean delivery | 56 days | Fever and dysnoea | Genital TB | Congenital TB | BAL = *Mycobacterium tuberculosis*  Mother= Genital TB diagnosed after birth = AFB positive | ATT  Initially mechanical ventilation | NA | Died |
| Samedi et al 2017 | Canada | 37-year  South Asian woman | At 24 weeks delivered a male child | 3 days post delivery | Seizures at 24 weeks of gestation | Disseminated TB  Miliary pulmonary TB and CNS TB | Congenital TB suspected | **Mother’s Placenta=** acid-fast bacilli. | ATT 2 months | Improved | Improved |
| Namani et al 2017 | *Kosovo* | 25-year-old Albanian woman  24 weeks of gestation after IVF | Preterm twins, 27 weeks of gestation | 14 days | Headache, fever, vomiting, coma | Multiple tuberculoma brain and TBM  Disseminated TB  Miliary pulmonary TB and CNS TB | NA | Urine culture= *Mycobacterium tuberculosis* | ATT 12 months | Improved | Both preterm twins died within 72 hours |
| Parmaksiz et al 2016 | Turkey | 30-year  Pregnant at 19 weeks via IVF | NA | 10 days | Fever, headache | Disseminated TB  Multi-drug resistant pulmonary TB  TBM | NA | Lung tissue biopsy = MDR-TB | Second line ATT | Improved | Pregnancy terminated at 19 weeks of gestation |
| Emiralioglu et al 2016 | Turkey | NA  3-month-old female infant  Born vaginally at 36 weeks' gestation | NA | In the 3rd week of life | Cough | Diagnosed retrospectively with genitourinary TB post-pregnancy | Congenital pulmonary TB, diagnosed post-birth  CNS TB | Gastric aspirate= *Mycobacterium tuberculosis* | ATT  12 months | NA | Full recovery |
| Hongbo and Li 2015 | China | 29-year  Pregnancy ongoing at 69 days gestation | NA | 5 days | Fever and shortness of breath | Miliary TB  CT chest= miliary lung lesions | NA | BAL PCR and culture = *Mycobacterium tuberculosis* | ATT | Improved | Patient had an abortion. |
| Zheng et al 2014 | China | 35-year | Born vaginally at 34 weeks' gestation | 3 days | Fever  Respiratory distress and hepatosplenomegaly | No active pulmonary TB | Congenital TB  Miliary TB  CT chest= miliary lung lesions | Endotracheal aspirate culture = *Mycobacterium tuberculosis* | ATT 8months  Mechanical ventilation | No active TB, latent infection likely | Improved |
| Ting-ting and Guo-zhong 2014 | China | 38 -year  Pregnancy ongoing at 29 weeks | NA | 14 days | Fever, headache, vision problems, diplopia, vomiting, history of tuberculosis | Disseminated TB  Miliary TB with CNS TB  CT chest= miliary lung lesions | NA | Positive T-SPOT.TB test for ESAT-6 and CFP-10 | ATT | Improved | NA |
| Mony et al 2014 | USA | 34-year  Emigrated from Nigeria | Female infant 27 5/7 weeks' gestation | 28 days | Cough and respiratory distress | Spinal TB diagnosed postpartum | Congenital pulmonary TB | Mother= Positive AFB smear from paraspinal collection  New born = Endotracheal aspirate culture = *Mycobacterium tuberculosis* | ATT | NA | Improved |
| Flibotte 2013  A report of five patients. | USA | 38-year | 31 weeks | 19 days | Late-onset sepsis, respiratory failure, seizures | Cold abscess, chronic ascites | Congenital TB  Right  upper lobe  infiltrate | *Post-mortem examination =*  *revealed multiple subpleural nodules and*  *smear= AFB* | NA | NA | Died |
|  |  | 38-year | 31 weeks | 19 days | Late-onset sepsis, respiratory failure, seizures | Cold abscess, chronic ascites | Congenital TB  Perihilar infiltrates  Pneumonia | Endometrial biopsy *culture grew M. africanum* | NA | NA | Improved |
|  |  | 30-year | 30 weeks | 24 days | Late-onset sepsis, respiratory failure | NA | Congenital pulmonary TB  Bilateral  pulmonary infiltrates | Endotracheal aspirate culture = *Mycobacterium tuberculosis*  Biopsy *culture grew Mycobacterium tuberculosis* | ATT | NA | Improved |
|  |  | 33-year | 35 weeks | 29 days | Fever and mild respiratory distress | Positive pre-IVF, calcified nodules and hilar lymph nodes | Congenital pulmonary TB  Miliary TB | Endotracheal and gastric aspirate culture = *Mycobacterium tuberculosis* | ATT | NA | Improved |
|  |  | 33-year | 35 weeks | 29 days | Fever and mild respiratory distress | Positive pre-IVF, calcified nodules and hilar lymph nodes | Congenital pulmonary TB  Pneumonia | Endotracheal and gastric aspirate culture = *Mycobacterium tuberculosis* | ATT | NA | Improved |
| Jacquemyn et al 2012 | *Belgium* | NA  Immigrant from Ghana | 13 weeks | 30 days | Persistent coughing, headache, fever and vaginal bleeding | Chest x-ray= miliary tuberculosis | None, fetus not infected | Blood cultures= *Mycobacterium tuberculosis* and placenta with acid-fast bacilli | ATT | Improved | Miscarriage at 13 weeks |
| Stuart et al 2009 | Australia | 29 -year  Bosnian Immigrant | 28 weeks | NA | Respiratory distress | Uterine granulomas 5 years prior; Positive IGRA Chest x-ray revealed old fibrotic lung changes | Congenital lymph node TB  Mediastinal lymphadenopathy with  extrinsic compression of trachea and bronchi | lymph node biopsy= Necrotizing granuloma  Lymph node and endotracheal aspirates  PCR and culture = *M. tuberculosis* | ATT  12 months | Uneventful recovery after the delivery | Improved |
| Altunhan et al 2009  (Twins) | Turkey | 34-year | *30 weeks* | 10 days | Respiratory distress | In postpartum diagnosed with tuberculous meningitis and multiple brain abscesses | Congenital pulmonary TB  X-RAY chest= Scattered infiltrates, consolidation, cavitation | CSF PCR= *M. tuberculosis*  Endometrial biopsy = necrotizing granulomatous endometritis  Endotracheal aspirates of newborn  = M. tuberculosis | ATT 2 months | Improved | Improved |
|  |  | 34-year | *30 weeks* | NA | Cough | In postpartum diagnosed with tuberculous meningitis and multiple brain abscesses | Congenital pulmonary TB  CT thorax= Scattered infiltrates, consolidation and calcifications | CSF PCR= M. tuberculosis  Endometrial biopsy = necrotizing granulomatous endometritis  Endotracheal aspirates of newborn  = M. tuberculosis | ATT 2 months | Improved | Improved |
| Chan et al 2003 | *Hong Kong* | 35-year | Newborn | 14 days | Newborn: Fever, pulmonary infiltrates, hepatosplenomegaly | CNS tuberculosis  Febrile encephalopathy with focal deficits | Congenital miliary tuberculosis | CSF= *Mycobacterium tuberculosis* DNA  Ziehl-Neelsen staining positive from gastric aspirate of newborn | ATT 12 months | Improved | Improved |
| Kissel et al 1999 | Germany | 37-year | NA | NA | Fever and headache | Miliary tuberculosis | NA | Histological examination showing granulomatous and necrotizing endometritis | ATT | Condition deteriorated | Abortion |
| Muller and Seelos 1994 | Germany | 30-year | NA | Since the 12th week of pregnancy | Fever, headache and vomiting | Miliary Tuberculosis,  Tuberculous Meningoencephalitis | NA | CSF PCR and smear= *Mycobacterium tuberculosis*  Vaginal fluid smear= *Mycobacterium tuberculosis* | ATT | Improved | Spontaneous abortion in the 23rd week of pregnancy |
| Addis et al 1988 | Scotland | 33-year | NA | Developed symptoms at 10 weeks | Flu-like symptoms, dry cough, dyspnea, vaginal bleeding, and fever | Miliary tuberculosis (*Mycobacterium bovis*)  X-ray chest = miliary lesions | NA | Urine culture positive for *Mycobacterium bovis* | ATT  6 months | Improved | Miscarriage at 14+ weeks |

ATT= Antituberculosis treatment; CNS= Central nervous system; DNA= Deoxyribonucleic acid; IGRA= Interferon Gamma Release Assay; IVF= In Vitro Fertilization; ICU = Intensive care unit; mNGS = Metagenomics Next Generation Sequencing; MDR= Multidrug resistant; MTP= Medical termination of pregnancy; MRS= Magnetic resonance spectrography; NA= Not available; NAAT= Nucleic Acid Amplification Testing; PCR= Polymerase chain reaction; TB = Tuberculosis

**Table-2: Details of clinical characteristics , treatment outcomes in patients with tuberculosis following In Vitro Fertilization (data were derived from cohort studies in case individual patients’ data was not available**

| **Reference** | **Country** | **Title of the study** | **Subjects (mother or newborn)** | **Summary of the study** |
| --- | --- | --- | --- | --- |
| Wei et al 2024 | China | Case-controlled study of tuberculosis in in-vitro fertilization-embryo transfer and natural pregnancy | Pregnant lady | Among 50 pregnant women with TB, 13 had conceived through IVF-ET. Significant differences between the IVF group and the natural conception group were observed. The IVF group tended to be older, had more frequent cases of fever, and showed a higher prevalence of severe TB forms, such as disseminated, extrapulmonary, and intracranial tuberculosis. Pregnancy termination was also more common in the IVF group compared to the natural conception group. |
| Zhang et al 2022 | China | Clinical characteristics in 26 children with congenital tuberculosis in Central Southern China: a retrospective study | Newborn | This study examined 26 children with congenital tuberculosis, born to mothers who conceived either naturally or through IVF. The median age of symptom onset was 25 days, with 73% occurring within four weeks. Most mothers (75%) were asymptomatic during pregnancy. Children born to IVF-ET mothers had more severe tuberculosis forms, such as tuberculous meningitis and liver tuberculosis, compared to those born naturally. Diagnosis was aided by an 84.2% positivity rate in T-SPOT.TB tests and imaging that revealed typical pulmonary lesions. Drug-resistant TB was rare, and the IVF-ET group had poorer outcomes, including higher mortality, liver injury, and developmental delays. |
| Xia et al 2022 | China | Association of in vitro fertilization with maternal and perinatal outcomes among pregnant women with active tuberculosis: A retrospective hospital-based cohort study | Pregnant lady | This study compared TB in pregnant or postpartum women who conceived through IVF with those who conceived naturally. Of 80 participants, 28 underwent IVF, with 67.9% experiencing fallopian tube obstruction. TB symptoms appeared earlier in the IVF group (13 vs. 22 weeks gestation). Although age and diagnostic delays were similar, the IVF group had more severe complications, including higher rates of vaginal bleeding (46.4% vs. 1.9%), maternal criticality (21.4% vs. 2.0%), miliary TB (89.3% vs. 13.5%), and TB meningitis (32.1% vs. 7.7%). |
| Wang et al 2022 | China | *Clinical analysis of pregnancy complicated with miliary tuberculosis* | Pregnant lady | This study analyzed 23 pregnant patients with miliary TB, including 12 who became pregnant through (VF-ET. Symptoms typically began at 13.96 weeks of gestation, with diagnosis taking an average of 33 days. Common symptoms were fever, dyspnea, cough, headache, and chest pain. Extrapulmonary TB was diagnosed in 10 patients, respiratory failure in 11, and acute respiratory distress syndrome (ARDS) in 9. All patients showed miliary nodules on chest HRCT. Six required mechanical ventilation, two needed ECMO, and one died. TB screening is crucial for women undergoing IVF-ET, with preventive treatment recommended for latent or untreated TB cases. |
| Dong et al 2022 | China | Analysis of clinical features and risk factors in pregnant women with Miliary Pulmonary Tuberculosis after in vitro fertilization embryo transfer | Pregnant lady | In this study, 75 pregnant women with TB were analyzed, including those who conceived naturally and those who underwent IVF-ET. Six patients in the IVF-ET group were diagnosed with miliary pulmonary tuberculosis. TB symptoms appeared earlier in the IVF-ET group, between 28 and 240 days after embryo transfer. All IVF-ET patients had fallopian tube obstruction, and 14.7% had a history of pulmonary or latent TB. The IVF-ET group experienced more severe symptoms, including fever (96%), dyspnea (46.7%), and vaginal bleeding (34.7%), with higher complication rates, such as respiratory failure (20%) and tuberculosis meningitis (17.3%). Imaging revealed multiple nodules (100%), ground-glass opacity (50%), and pleural effusion (50%). Despite treatment with first-line anti-TB therapy, only one baby survived in the IVF-ET group, with most pregnancies resulting in spontaneous abortion (33.3%), artificial termination (16.7%), or preterm delivery with neonatal death (16.7%). The natural pregnancy group had fewer severe symptoms and better fetal outcomes. |
| QIN et al 2021 | China | Analysis of clinical characteristics and prognosis of 21 pregnant women complicated with tuberculosis after in vitro fertilization-embryo transfer | Pregnant lady | This study of 30 TB patients following IVF-ET revealed significant complications. Miliary TB was seen in 66.7%, tuberculous meningitis in 33.3%, and tuberculous peritonitis in 13.3%. Respiratory failure affected 3 patients, with ICU admission required. Miscarriage or preterm labor occurred in 60%, with 15 patients undergoing elective abortion. Despite treatment, one patient remained comatose from meningitis. IVF-ET patients experienced more severe TB complications compared to natural pregnancies, highlighting a more challenging TB course in this group. |
| Huang et al 2021 | China | The Clinical Features and Prognosis Analysis of Pregnant Women with Tuberculosis After in Vitro Fertilization and Embryo Transfer and Natural Fertilization | Pregnant lady | In a preprint cohort study, 95 cases were analyzed, consisting of 24 cases of tuberculosis following IVF and 71 cases of tuberculosis in natural pregnancies. Pregnant women with tuberculosis after in vitro fertilization and embryo transfer experienced more severe forms of the disease, with increased systemic symptoms, a higher incidence of drug-induced liver injury, and elevated rates of spontaneous and inevitable abortions compared to those with natural pregnancies. |
| Gai et al 2021 | China | Acute miliary tuberculosis in pregnancy after in vitro fertilization and embryo transfer: a report of seven cases | Pregnant lady | This study examined seven women who developed TB following IVF-ET. All patients, aged 28-35 years, were diagnosed with miliary TB during pregnancy, and two had TB meningitis. Six patients had old TB lesions on chest X-rays before IVF-ET but did not receive prior anti-TB treatment. Despite receiving anti-TB therapy after diagnosis, pregnancy outcomes were poor, with four spontaneous and three induced abortions. These findings suggest that TB reactivation is severe in post-IVF-ET pregnancies, highlighting the importance of pre-IVF TB screening and treatment to improve outcomes. |
| Du et al 2021 | China | Clinical characteristics and post-discharge follow-up analyses of 10 infants with congenital tuberculosis: A retrospective observational study | Newborn | In this study, two out of ten infants (20%) with congenital TB were born following IVF. Both mothers had a prior history of TB but were asymptomatic during pregnancy. TB reactivation occurred after the infants' diagnosis. Both infants presented with fever, and chest CT scans revealed multiple pulmonary nodules. Anti-TB treatment, including isoniazid (INH) and rifampicin (RIF), was initiated. One infant (50%) experienced neurological complications, including seizures, requiring physical therapy. Despite these issues, both infants survived, emphasizing the need for TB monitoring in IVF pregnancies, especially in cases with a maternal history of TB. |
| Yu et al 2019 | China | Clinical analysis of 15 cases of tuberculous meningitis after in vitro fertilization and embryo transplantation | Pregnant lady | This study retrospectively analyzed 23 cases of TB meningitis in pregnant women following IVF-ET. Common symptoms included fever, with 60.9% experiencing high fever. Miliary TB was present in 65.2%, and pleurisy in 34.8%. Brain MRI abnormalities were found in 78.3%. T-SPOT.TB was positive in 60.9%, but cerebrospinal fluid cultures were positive in only 26.1%. After treatment, 91.3% of patients were cured, though two died. Among 13 infants, one died of congenital TB. The study highlights the prevalence of severe TB forms and the need for prolonged treatment in post-IVF TB meningitis cases. |
| Ye et al 2019 | China | Characteristics of miliary tuberculosis in pregnant women after in vitro fertilization and embryo transfer | Pregnant lady | This study examined six cases of miliary TB in pregnant women following IVF-ET. The patients, aged 29–39 years, had symptoms onset between 5 and 26 weeks of gestation, with diagnosis delays of 10 to 32 days. Fever, dyspnea, and cough were common symptoms. All patients had elevated inflammatory markers, and chest CT scans showed diffuse miliary infiltrations. One patient was diagnosed with TB meningitis. Despite receiving anti-TB treatment, all pregnancies ended in fetal loss, with three patients experiencing spontaneous abortion and the remaining three undergoing artificial pregnancy termination. |

**Refences**

1. Wei JL, Zhang L, Xu YL, Gan W, Qi M, Fu XW, Li X. Case-controlled study of tuberculosis in in-vitro fertilisation-embryo transfer and natural pregnancy. BMC Pregnancy Childbirth. 2024 Jan 23;24(1):77. doi: 10.1186/s12884-024-06260-1.
2. Zhang F, Zhang XF, Zhou HY. Clinical characteristics in 26 children with congenital tuberculosis in Central Southern China: a retrospective study. Paediatr Int Child Health. 2022 Aug-Nov;42(3-4):127-132. doi: 10.1080/20469047.2023.2246006.
3. Xia L, Mijiti P, Liu XH, Hu ZD, Fan XY, Lu SH. Association of *in vitro* fertilization with maternal and perinatal outcomes among pregnant women with active tuberculosis: A retrospective hospital-based cohort study. Front Public Health. 2022 Oct 17;10:1021998. doi: 10.3389/fpubh.2022.1021998.
4. Wang K, Ren D, Qiu Z, Li W. Clinical analysis of pregnancy complicated with miliary tuberculosis. Ann Med. 2022 Dec;54(1):71-79. doi: 10.1080/07853890.2021.2018485.
5. Dong S, Zhou R, Peng E, He R. Analysis of Clinical Features and Risk Factors in Pregnant Women With Miliary Pulmonary Tuberculosis After *In Vitro* Fertilization Embryo Transfer. Front Cell Infect Microbiol. 2022 Jul 11;12:885865. doi: 10.3389/fcimb.2022.885865.
6. QIN Y, CAI QS, BAO ZJ. Analysis of clinical characteristics and prognosis of 21 pregnant women complicated with tuberculosis after in vitro fertilization-embryo transfer. Chinese Journal of Antituberculosis. 2021 Jul 10;43(7): 682-688. doi: [10.3969/j.issn.1000-6621.2021.07.008](https://doi.org/10.3969/j.issn.1000-6621.2021.07.008)
7. Huang W, Li T, Liu P, Liu J, Dong L, Xi X, Liu X, Xia L, Lu S. The Clinical Features and Prognosis Analysis of Pregnant Women with Tuberculosis After in Vitro Fertilization and Embryo Transfer and Natural Fertilization. Research Square Preprint. *DOI: https://doi.org/10.21203/rs.3.rs-918630/v1*
8. Gai X, Chi H, Cao W, Zeng L, Chen L, Zhang W, Song D, Wang Y, Liu P, Li R, Sun Y. Acute miliary tuberculosis in pregnancy after in vitro fertilization and embryo transfer: a report of seven cases. BMC Infect Dis. 2021 Sep 6;21(1):913. doi: 10.1186/s12879-021-06564-z.
9. Du J, Dong S, Jia S, Zhang Q, Hei M. Clinical characteristics and post-discharge follow-up analyses of 10 infants with congenital tuberculosis: A retrospective observational study. Pediatr Investig. 2021 Jun 18;5(2):86-93. doi: 10.1002/ped4.12266.
10. Yu CH, LI QS, FAN LC, ZHU YJ, ZHANG L, ZHOU MF, LYU YQ, YU ZY. Clinical analysis of 15 cases of tuberculous meningitis after in vitro fertilization and embryo transplantation. Journal of Tuberculosis and Lung Disease. 2019;8(4): 280-284. doi: [10.3969/j.issn.2095-3755.2019.04.009110000](https://doi.org/10.3969/j.issn.2095-3755.2019.04.009110000)
11. Ye R, Wang C, Zhao L, Wu X, Gao Y, Liu H. Characteristics of miliary tuberculosis in pregnant women after in vitro fertilisation and embryo transfer. Int J Tuberc Lung Dis. 2019 Feb 1;23(2):136-139. doi: 10.5588/ijtld.18.0223.
